# Cost-effectiveness of antigen testing for ending COVID-19 isolation

**DOI:** 10.1101/2022.03.21.22272687

**Authors:** Sigal Maya, James G. Kahn

## Abstract

**Background:** The Omicron variant of SARS-CoV-2 led to a steep rise in transmissions. Recently, as public tolerance for isolation abated, CDC guidance on duration of at-home isolation of COVID-19 cases was shortened to five days if no symptoms, with no lab test requirement, despite more cautious approaches advocated by other federal experts.

**Methods:** We conducted a decision tree analysis of alternative protocols for ending COVID-19 isolation, estimating net costs (direct and productivity), secondary infections, and incremental cost-effectiveness ratios. Sensitivity analyses assessed the impact of input uncertainty.

**Results:** Per 100 individuals, five-day isolation had 23 predicted secondary infections and a net cost of $33,000. Symptom check on day five (CDC guidance) yielded a 23% decrease in secondary infections (to 17.8), with a net cost of $45,000. Antigen testing on day six yielded 2.9 secondary infections and $63,000 in net costs. This protocol, compared to the next best protocol of antigen testing on day five of a maximum eight-day isolation, cost an additional $1,300 per secondary infection averted. Antigen or polymerase chain reaction testing on day five were dominated (more expensive and less effective) versus antigen testing on day six. Results were qualitatively robust to uncertainty in key inputs.

**Conclusions:** A six-day isolation with antigen testing to confirm the absence of contagious virus appears the most effective and cost-effective de-isolation protocol to shorten at-home isolation of individuals with COVID-19.

## Introduction

The SARS-CoV-2 B.1.1.529 (Omicron) variant was designated a variant of concern by the World Health Organization in November 2021, as it had several mutations that are suspected to impact its transmissibility and disease severity.[1, 2] In late December, in the US, where vaccination coverage is above 60%,[3] public health guidance from the Centers for Disease Control and Prevention (CDC) regarding isolation of COVID-19 cases were relaxed. The recommended duration of isolation was decreased from ten to five days, with no laboratory testing required to end isolation.[4] Individuals were asked to evaluate their symptoms on day five to determine whether to continue isolating for the full ten days. This guidance was received with skepticism[5-7] given the understanding that Omicron, constituting 95% of COVID-19 cases in the US,[8] was potentially more transmissible than earlier variants and less susceptible to vaccines.[1, 2] Shortages of rapid antigen tests in the US,[7, 9] economic losses associated with extended periods of isolation,[10] and the psychological effects of longer at-home isolation durations[11] were suggested as possible reasonings behind the updated guidance. However, fully elaborated scientific evidence supporting the decision was lacking.[12]

While quantitative studies on the viral kinetics and pathophysiology of the Omicron variant are underway, decision makers must offer timely guidance that balances public health and economic considerations in their COVID-19 isolation recommendations. Antigen testing could offer benefits over using symptom status as a marker of infectivity given the high rate of asymptomatic COVID-19 infections.[13] We aimed to evaluate the trade-offs between costs (including lost productivity) and secondary infections averted when adopting different protocols to end COVID-19 isolation in order to provide an evidence-base for such decisions.

## Materials and Methods

### Model design

We modeled six different protocols for ending COVID-19 isolation. Using a customized decision tree template, we compared the number of secondary COVID-19 infections that occurred when individuals followed each of these different protocols. We adopted a societal perspective and a two-week time horizon to capture all costs and secondary infections. The cohort consisted of 100 individuals in the US who had COVID-19 (confirmed by PCR and/or antigen test) and were on the fifth day of isolation. We modeled only individuals with asymptomatic or mild COVID-19; those with more severe disease would be hospitalized rather than isolating at home and therefore were not included.

### De-isolation protocols

Interventions were selected to demonstrate current policy options as well as alternatives that might reduce transmissions while also shortening isolation duration. While not exhaustive, these protocols represent a variety of options that might warrant further evaluation. In all strategies, individuals leaving isolation were assumed to follow best practices for infection prevention, which at the time of the analysis included mask wearing. Individuals could leave their home the day after their isolation ended (i.e., for a five-day isolation, they spent five full days at home and could leave on day six if cleared).

#### Five-day isolation

Person with confirmed COVID-19 stays at home for five days, then can leave without any further consideration.

#### Ten-day isolation with symptom check on day five (i.e., the CDC guidance)

Person with confirmed COVID-19 stays at home for five days. On day five, they review their symptoms. Those who were asymptomatic or fever free for 24 hours can end isolation, while those with persisting symptoms continue to isolate until day ten.

#### Ten-day isolation with antigen test on day five

Person with confirmed COVID-19 stays at home for five days. On day five, they perform a rapid antigen test. Those who test negative can end isolation while those who test positive continue to isolate until day ten.

#### Ten-day isolation with PCR test on day five

Person with confirmed COVID-19 stays at home for five days. On day five, they conduct a PCR test. Those who test negative can end isolation while those who test positive continue to isolate until day ten. We assumed results are obtained within 24 hours.

#### Ten-day isolation with antigen test on day six

Person with confirmed COVID-19 stays at home for six days. On day six, they perform a rapid antigen test. Those who test negative can end isolation while those who test positive continue to isolate until day ten.

#### Eight-day isolation with antigen test on day five

Person with confirmed COVID-19 stays at home for five days. On day five, they perform a rapid antigen test. Those who test negative can end isolation while those who test positive continue to isolate until day eight instead of day ten (no re-test is done).

### Key assumptions

We assumed that no one in the cohort was SARS-CoV-2-naïve (i.e., all had begun isolation based on true-positive test results). As such, individuals were either still carrying contagious virus or had cleared all viable virus. We defined a frontloaded distribution for infectivity over ten days following symptom onset (or positive test, if asymptomatic), based on empirical data on culture-positivity of patient samples.[14-17] For those remaining in isolation after day five, we assumed imperfect isolation effectiveness such that continued isolation led to a 95% reduction in the risk of transmission. Finally, for illustration purposes, we assumed 100% testing coverage (i.e., everyone had access to the tests necessary). This was varied in sensitivity analyses.

### Model inputs

We used data specific to the Omicron variant when available to parameterize the model. Otherwise, we used data generated during the wildtype (Alpha) and B.1.617.2 (Delta) variant waves. Key model inputs are presented in Table 1, with uncertainty ranges and sources.

**Table 1.**
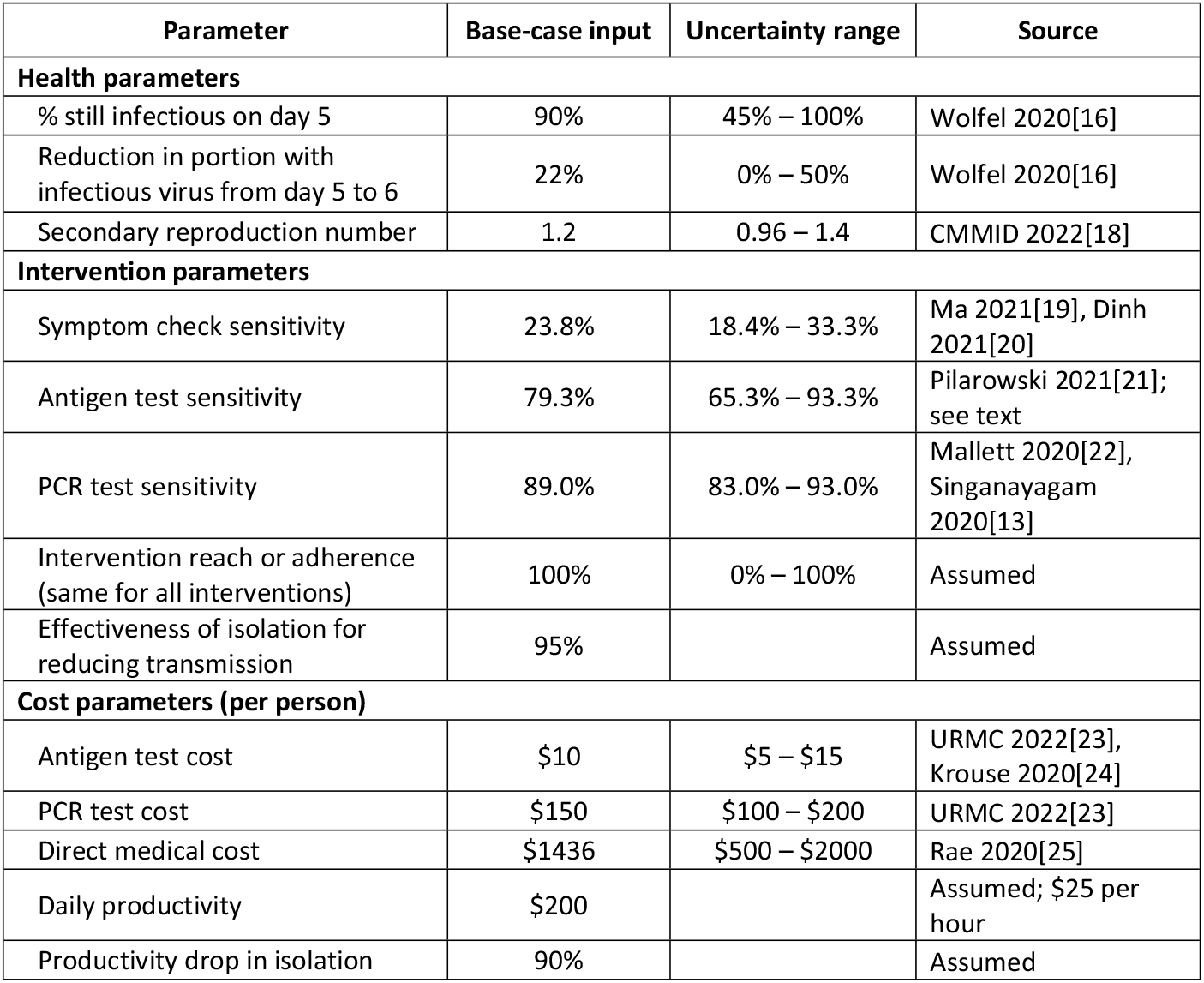
Model parameters, uncertainty ranges, and sources.

| Parameter | Base-case input | Uncertainty range | Source |
| --- | --- | --- | --- |
| **Health parameters** |  |  |  |
| % still infectious on day 5 | 90% | 45% – 100% | Wolfel 2020[16] |
| Reduction in portion with infectious virus from day 5 to 6 | 22% | 0% – 50% | Wolfel 2020[16] |
| Secondary reproduction number | 1.2 | 0.96 – 1.4 | CMMID 2022[18] |
| **Intervention parameters** |  |  |  |
| Symptom check sensitivity | 23.8% | 18.4% – 33.3% | Ma 2021[19], Dinh 2021[20] |
| Antigen test sensitivity | 79.3% | 65.3% – 93.3% | Pilarowski 2021[21]; see text |
| PCR test sensitivity | 89.0% | 83.0% – 93.0% | Mallett 2020[22], Singanayagam 2020[13] |
| Intervention reach or adherence (same for all interventions) | 100% | 0% – 100% | Assumed |
| Effectiveness of isolation for reducing transmission | 95% |  | Assumed |
| **Cost parameters (per person)** |  |  |  |
| Antigen test cost | \$10 | \$5 – \$15 | URMC 2022[23], Krouse 2020[24] |
| PCR test cost | \$150 | \$100 – \$200 | URMC 2022[23] |
| Direct medical cost | \$1436 | \$500 – \$2000 | Rae 2020[25] |
| Daily productivity | \$200 | | Assumed; \$25 per hour |
| Productivity drop in isolation | 90% |  | Assumed |

#### Health inputs

The probability of carrying viable SARS-CoV-2 was 90% on day five and 70% on day six, quickly dropping to zero by day ten from the start of isolation, based on studies of previous variants.[13-17] The effective secondary reproduction number (R_eff_), which implicitly accounts for infection prevention measures that were in place such as mask wearing, was used to calculate the number of secondary infections that occur per index case. As of January 16^th^, 2022, R_eff_ was estimated as 1.2 in the US.[18] We adjusted this value based on the probability of carrying infectious virus on each day of infection to reflect the reduced transmissibility five days after the start of isolation; the “residual R” was 0.26 over days six to ten if isolation was discontinued (See S1 Appendix for calculations). Similar calculations were made to adjust R_eff_ for de-isolation protocols requiring longer isolation periods.

#### Test performance

Forty percent of relevant COVID-19 cases were asymptomatic,[19] and of those who develop symptoms, 60% had symptom resolution by day five post-symptom onset,[20] yielding approximately 24% sensitivity for the symptom check protocol. Previous studies among mildly symptomatic and asymptomatic individuals showed antigen tests had over 93% sensitivity for viral loads high enough to be transmissible.[21] We reduced this value by 15% to account for the suspected reduction in sensitivity for the Omicron variant,[1] which resulted in approximately 80% antigen test sensitivity. PCR tests had 89% sensitivity.[22]

#### Cost inputs

Costs were calculated from a societal perspective and in 2022 US dollars. A rapid antigen test cost $10, while a PCR test cost $150.[23, 24, 26] Productivity loss due to isolation was $900 over five days, assuming a 90% decrease in productivity and $200 per day. This is likely an overestimate of productivity loss since many individuals with asymptomatic COVID-19 isolating at home can continue working remotely with no or minimal loss in productivity. We therefore calculated base-case outputs both with and without productivity loss. Direct medical costs incurred for secondary infections were $1436 on average; this accounted for varying costs for different disease severity levels (e.g., $0 if asymptomatic or no healthcare is sought vs. $61,000 if ICU admission is required; see S1 Appendix).[25, 27] We assumed all medical costs were incurred in year one and did not require discounting.

### Model outputs and sensitivity analyses

We compared the number of secondary infections, societal net costs, and, when appropriate, incremental cost-effectiveness ratios (ICERs) given different de-isolation protocols. De-isolation protocols that led to fewer net costs and fewer secondary infections than their comparator were dominant; no ICERs were calculated.

We conducted deterministic and probabilistic sensitivity analyses to assess uncertainty in key inputs in Table 1. Since test availability and protocol adherence was arbitrarily set as 100%, these two inputs were not included in one-way and multivariate (Monte Carlo) sensitivity analyses. Instead, we performed threshold analyses and two-way sensitivity analyses (where inputs were varied two at a time) to determine minimum necessary adherence and test availability for de-isolation protocols to be effective. When test availability was not 100%, individuals left isolation after the day on which they would have otherwise taken a test.

Additionally, we simulated three separate risk scenarios to evaluate how the environment individuals are re-entering upon ending isolation would affect outcomes. The change in the risk of infection due to varying vaccination rate, mask-wearing[3, 28-30], and number of contacts[31] from base-case were used to adjust the transmission rate, R_eff_ (see S1 Appendix). A *low-risk scenario* was defined representing the infected individual re-joining the household where everyone was fully vaccinated and continued to wear masks for the next five days (R_eff_=0.35, residual R after de-isolation=0.07). A *medium-risk scenario* reflected individuals starting to see few non-household members, all of whom were fully vaccinated, but mask-wearing was inconsistent (R_eff_=1.11, residual R=0.24). Finally, a *high-risk scenario* was defined in which both vaccination and mask-wearing was inconsistent, and a greater number of social contacts were occurring (e.g., going to the movies, eating at restaurants, attending school; R_eff_=3.78, residual R=0.81). We did not conduct multivariate sensitivity analyses on the scenarios.

### Statistical analysis

The model was built in Excel® (Office 365, Microsoft Corporation) and sensitivity analyses were conducted using @RISK® (version 8.2, Palisade Corporation). The decision tree and all data are available upon request.

## Results

Base-case results from a societal perspective (i.e., including productivity loss due to isolation) are presented in Table 2; all outcomes are given per 100 individuals. Ending isolation at day five without further testing led to 23.0 secondary infections and $33,100 in direct medical costs. Symptom check at day five (17.8 secondary infections) reduced transmissions by 23% with a $11,900 increase in net costs; the ICER was $2,282 per secondary infection averted.

**Table 2.**
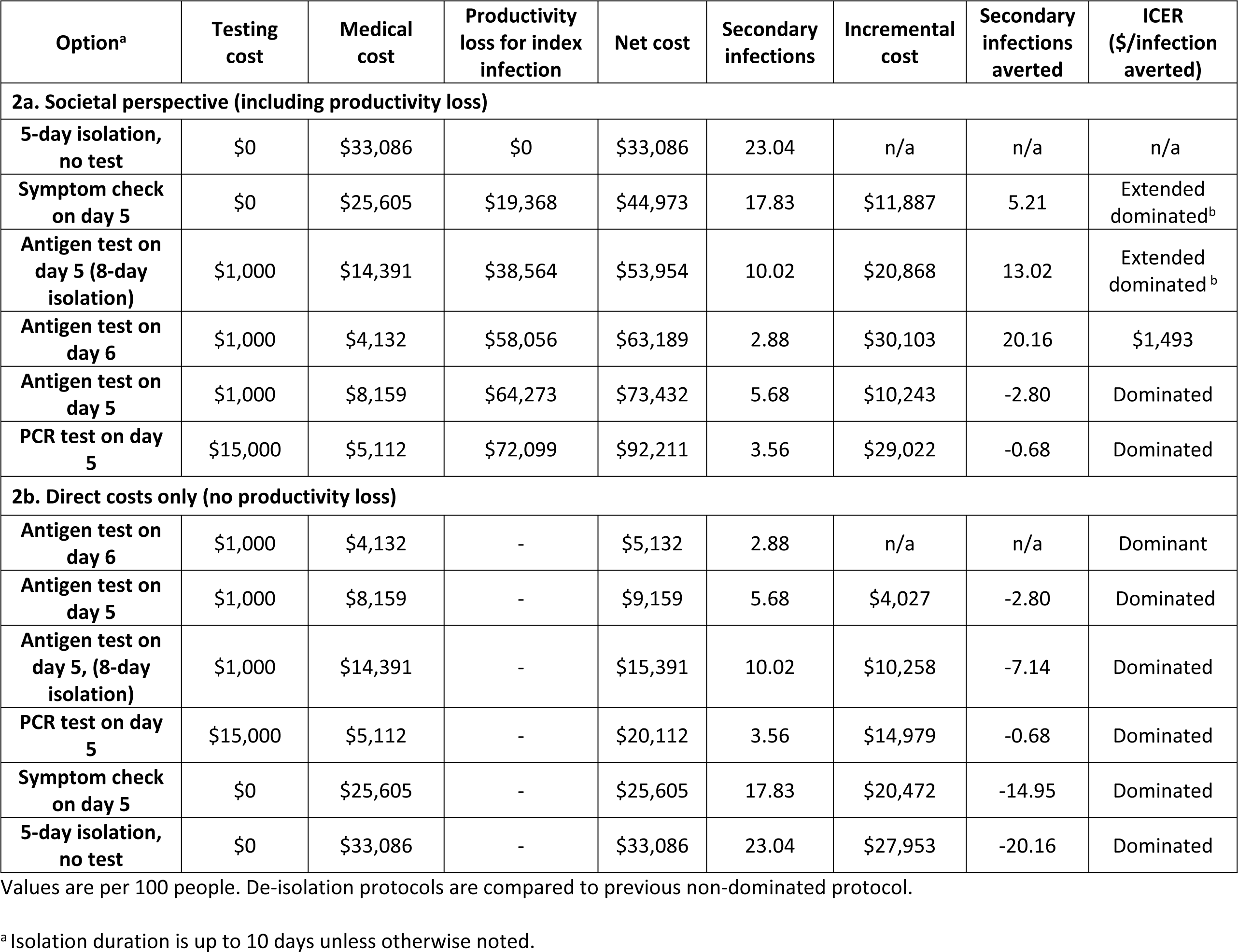
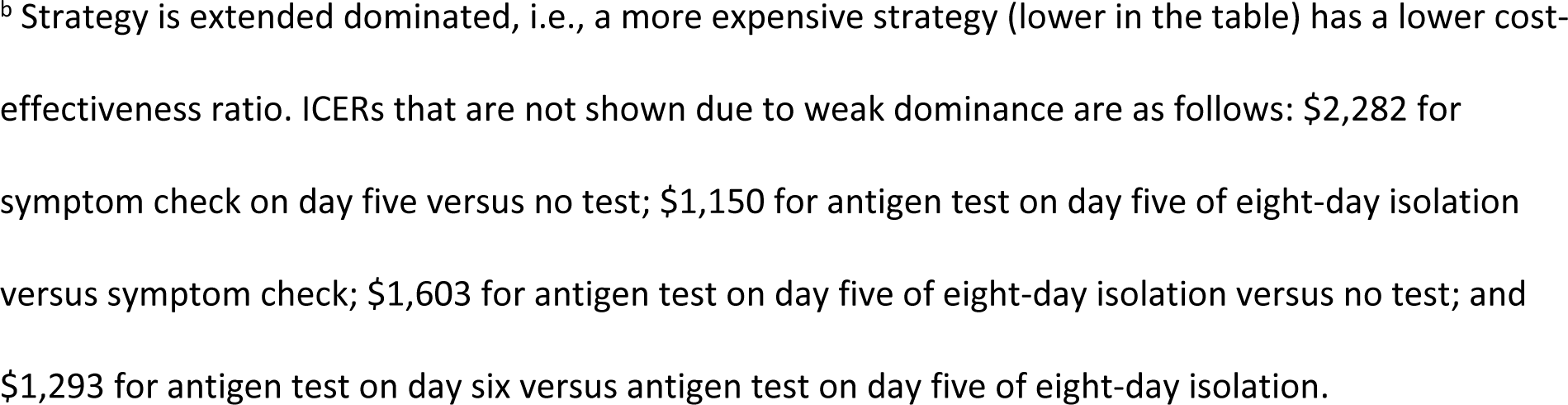
Base-case results from (a) societal perspective (including productivity loss) and (b) with direct costs only (no productivity loss).

Antigen testing on day five of an eight-day isolation period cost an additional $1,150 per secondary infection averted compared with symptom check. This drop in the ICER represents extended dominance[32] over the symptom check. The ICER for day five antigen test versus no test was $1,603.

The most cost-effective de-isolation protocol was performing an antigen test on day six of a ten-day isolation period. This protocol led to $63,200 in net costs and 2.9 secondary infections, yielding an ICER of $1,293 per secondary infection averted versus an antigen test on day five of an eight-day isolation, again representing extended dominance. Both antigen and PCR testing on day five were dominated by antigen testing on day six; they led to greater net costs and more secondary infections.

If productivity losses were omitted, leaving just direct costs, antigen testing on day six was strictly dominant (i.e., lowest net cost and fewest secondary infections) over all other de-isolation protocols (Table 2).

### Sensitivity analyses

In one-way sensitivity analyses where key inputs were varied one at a time, antigen testing on day five prevented between 51-85% secondary infections over symptom check, depending primarily on antigen test sensitivity for transmissible viral loads. Secondary infections prevented with an antigen test on day six versus day five was mostly related to the relative reduction in viable viral load from day five to six and varied between 35-67%. Antigen test on day six, compared to the next most cost-effective option (antigen test on day five of eight-day isolation) prevented between 3.6 and 8.0 secondary infections. This value was most sensitive to the probability of having transmissible virus on day five (Figure 1).

**Fig 1.**
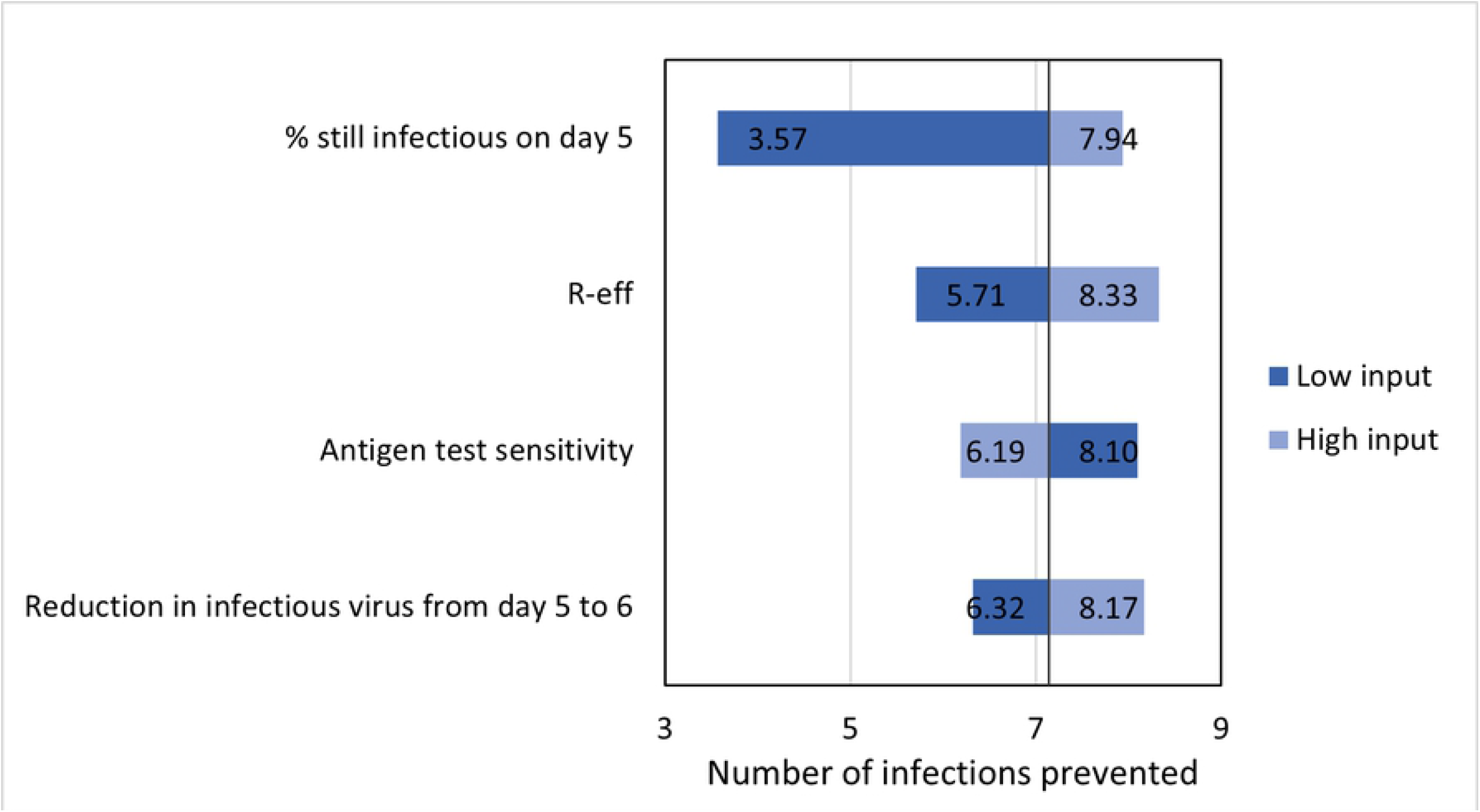
One-way sensitivity analyses on the number of secondary infections averted with antigen test on day six versus next most cost-effective strategy (eight-day isolation with antigen test on day five). Base-case output is 7.14.

Probabilistic Monte Carlo analyses showed that antigen testing on day six was either dominant or cost-effective with ICERs up to $3,816 per secondary infection averted, given varying inputs. This outcome was most sensitive to uncertainty in the probability of a persistent high viral load, followed by the community transmission rate and the direct medical cost per COVID-19 infection. Antigen test on day six always prevented more secondary infections than antigen test on day five of eight-day isolation but had a nearly 90% probability of having greater net costs (Figure 2).

**Fig 2.**
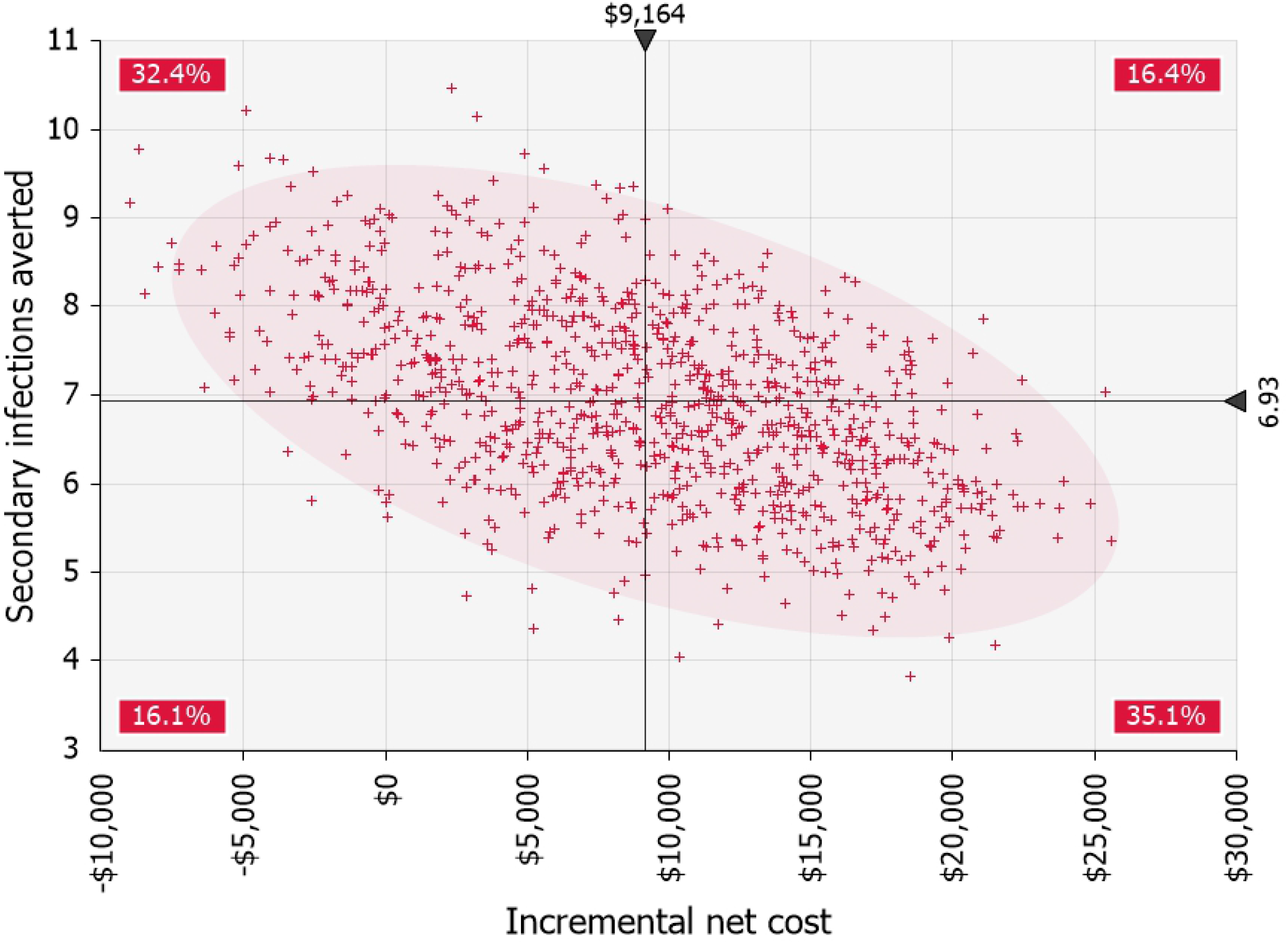
Simulated incremental costs and secondary infections averted with antigen testing on day six of isolation versus next most cost-effective strategy (eight-day isolation with antigen test on day five). 1000 iterations. Shading represents 95% confidence area for results. Percentages are the probabilities of the result being in each of the quadrants.

In all three of the risk scenarios considered, antigen testing on day six remained the optimal de-isolation protocol (Table 3). Both the low- and medium-risk scenario results followed base-case findings: symptom check at day five, antigen test on day five of eight-day isolation, and antigen test on day six were all cost-effective with ICERs increasing as transmission risk decreases. The ICER for antigen testing on day six was $8,050 and $1,500 per secondary infection averted in the low- and medium-risk scenarios, respectively. In the high-risk scenario, antigen testing on day six was strictly dominant, leading to $72,000 in net costs and 9.1 secondary infections, as opposed to $100,000 net costs and 72.6 secondary infections with a symptom check on day five.

**Table 3.**
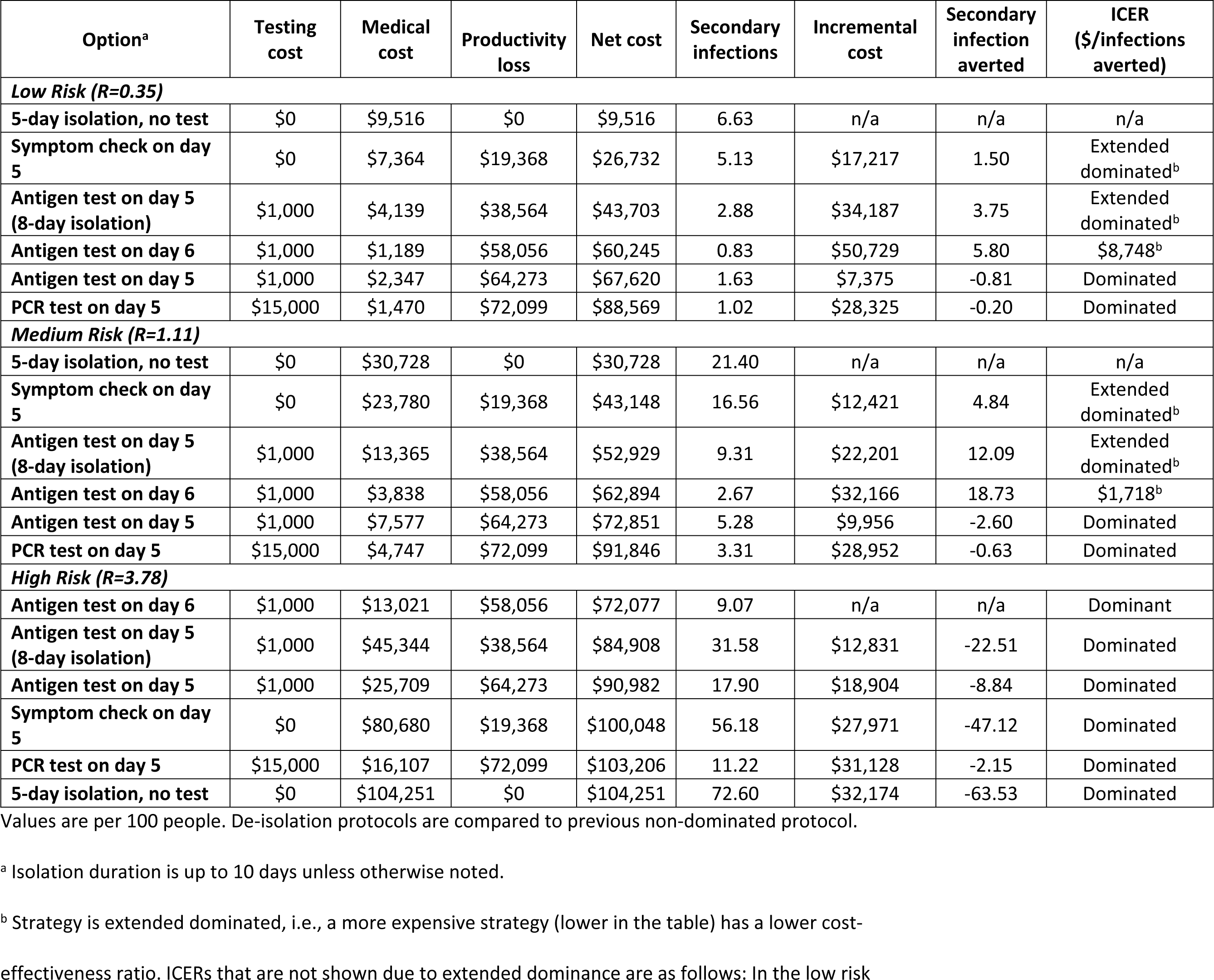
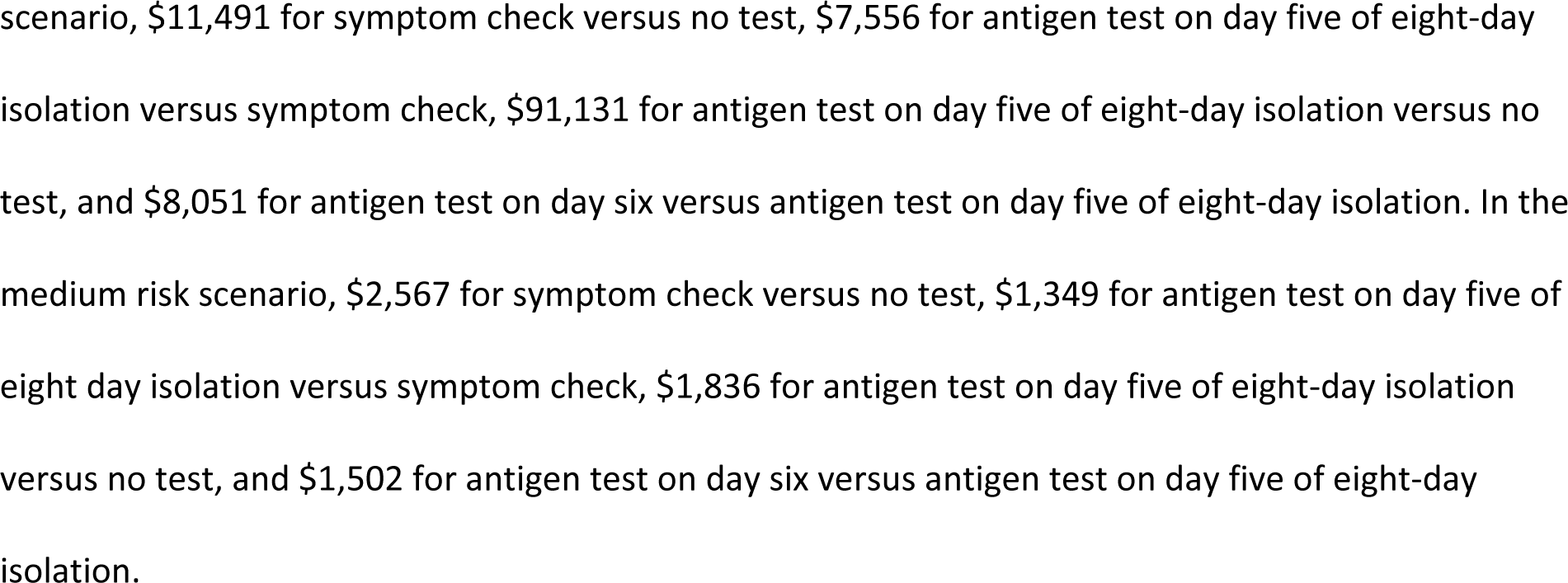
Analyses of different risk scenarios. Values per 100 people.

Under the base-case assumption of 100% adherence to a symptom check protocol, at least 30% antigen test availability (i.e., 30% of those in isolation can access a test) was necessary for antigen testing to prevent more secondary infections than the symptom check if done on day five. Similarly, PCR tests had greater benefit than symptom check when test availability was greater than approximately 27%. In a two-way sensitivity analysis, if symptom check adherence was below 70%, then <20% antigen test or <19% PCR test availability was sufficient for these tests to prevent a greater number of transmissions than the symptom check protocol on day five. Notably, antigen testing on day six prevented more secondary infections than symptom check on day five even when test availability was as low as 1%, due to the added day of isolation.

## Discussion

We compared health and cost outcomes associated with different de-isolation protocols to end COVID-19 isolation for those with confirmed asymptomatic or mild COVID-19. All ICERs we calculated had favorable cost-effectiveness ratios. We found that while symptom check without testing on day five of isolation did reduce secondary transmissions after de-isolation by 23% compared to no testing, it still led to nearly 18 secondary infections per 100 individuals and had the least favorable cost-effectiveness ratio due to high medical costs for secondary infections. The most cost-effective protocol was to remain in isolation through day six and then perform an antigen test, which dominated both antigen testing and PCR testing on day five. Antigen testing on day six led to an overall 87% decrease in secondary infections compared to no testing and cost an additional $1,300 per secondary infection averted compared to the next best option.

Notably, threshold analysis on antigen test availability suggested that the benefit of antigen testing on day six might be primarily due to the extra day of isolation (during which the probability of still carrying infectious virus quickly begins to drop), rather than the ability of the test to identify those who still might have a transmissible viral load. This insight warrants further evaluation using emerging data on the viral dynamics of Omicron. Moreover, antigen testing on day six was associated with lower productivity loss than antigen testing on day five; even though everyone remained in isolation for one more day, more individuals were cleared for de-isolation on day six than would have been on day five. The four days gained by this portion of index cases offset the extra day lost by everyone. Workforce shortages have been an important adverse effect of COVID-19 isolation.[33-37] Antigen testing on day six generated both health and economic benefits; it minimized post-isolation transmissions while allowing individuals to return to work sooner on average.

By modeling different risk scenarios, we demonstrated that the de-isolation environment had a considerable impact on the cost-effectiveness of testing strategies. Regardless of the risk scenario, the optimal protocol remained antigen testing on day six, which became dominant over other protocols in high-risk situations and remained cost-effective, although cost-effectiveness was less favorable in low-risk situations. Nevertheless, these findings suggest that much like other public health policy decisions throughout the pandemic, de-isolation guidelines must evolve as the context of the pandemic shifts. For example, potential new surges may call for more stringent policies with longer minimum isolation and more sensitive tests, while declining transmissions may allow more lenient approaches. It is plausible that the CDC has reached this same conclusion and proposed a symptom check rather than an antigen test because of an expectation that transmissions would subside in the weeks following the announcement of the new guidance. While reasonable at first glance, this could be a risky approach; loosening infection prevention measures may have prevented the expected drop in transmissions, leading instead to a quick rise in cases that would have prohibited the loosened guidance being put in place to begin with. Indeed, our modeling of the CDC guidance resulted in a substantial number of secondary COVID-19 cases given the transmission rate at the time the guidance was issued, some of which were avoided with a different de-isolation approach.

Evidence from earlier SARS-CoV-2 variants suggest that transmissibility of the virus peaks by approximately the fifth day from symptom onset, and swiftly drops afterward.[13-17, 38] However, the risk of further transmission after day five is not zero, and it is highly dependent on health behavior following de-isolation (e.g., continuing to wear masks, limiting the number of social contacts etc.) Given the high probability of asymptomatic infection and the possibility of short-lived symptoms, a symptom check to end isolation on the fifth day does not substantially reduce the risk of further transmission. Antigen tests, on the other hand, allow a more accurate measure of ongoing risk. There is now evidence that these rapid tests have good sensitivity for detecting high viral loads that are most likely to be transmissible.[17, 21, 38, 39] As such, antigen tests are an important public health tool that can help mitigate the health harms of the COVID-19 pandemic, and should be incorporated into public health responses as resources allow.

### Limitations

This study had important limitations, especially regarding uncertainty in key inputs such as the viral kinetics of SARS-CoV-2 and sensitivity of antigen tests. First, we distributed R_eff_ over the 10 days following COVID-19 confirmation, but a portion of transmissions occur prior to the index case learning their COVID-19 status and entering isolation. As such, we have overestimated the number of secondary infections in our model and underestimated ICER values. Decreasing the residual R would increase ICERs but given favorable ICERs even in the low-risk scenario, we believe the implications of our findings would not be affected. More importantly, given the novelty of the Omicron variant, we had to rely on studies of prior variants for these two important factors. While variance in neither of these inputs changed cost-effectiveness results qualitatively, they did have an impact on the number of further transmissions after isolation and thus the level of cost-effectiveness. Additional studies on the viral kinetics of the Omicron variant are necessary to refine these estimates.

## Conclusions

The Omicron variant of SARS-CoV-2 presents a new threat to public health due to its high transmissibility and potential ability to evade vaccine-induced immunity. Cost-effectiveness analyses can help decision makers assess the trade-offs between the economic disadvantages and health risks of adopting different COVID-19 de-isolation guidance. Using a decision tree model and Omicron-specific data when available, we found that ending isolation in five days given a negative symptom check left substantial risk of transmission and was not the most cost-effective strategy even when high productivity losses of longer isolation were accounted for. Instead, our findings suggest a baseline isolation duration of six days, at which time an antigen test, if available, can be conducted to confirm that the individual no longer carries transmissible SARS-CoV-2.

## Data Availability

All relevant data are within the manuscript and its Supporting Information files.

## Acknowledgements

The authors thank Dr. Elliot Marseille for review.

## Supporting Information

**S1 Appendix. Medical costs, transmission rates, infectivity**.

